# Skin-to-skin holding in relation to white matter connectivity in infants born preterm

**DOI:** 10.1101/2025.03.21.25324424

**Authors:** Katherine E. Travis, Molly F Lazarus, Melissa Scala, Virginia A. Marchman, Lisa Bruckert, Rocio Velasco Poblaciones, Sarah Dubner, Heidi M. Feldman

## Abstract

**Background and Objectives:** Preterm birth is associated with altered white matter development and long-term neurodevelopmental impairments. Skin-to-skin care (kangaroo care) has well-documented benefits for physiological stability and bonding, but its association with neonatal brain structure remains unclear. This study explored the association between in-hospital skin-to-skin care and neonatal white matter microstructure in frontal and limbic pathways that are linked to stress regulation and socio-emotional development, processes potentially influenced by affective touch during skin-to-skin care.

**Methods:** This retrospective study analyzed electronic medical records and diffusion MRI data collected from 86 preterm infants (<32 weeks gestational age) in a single NICU. Skin-to-skin care exposure was quantified as total duration (minutes/instance) and rate (minutes/day) of sessions. Diffusion MRI scans obtained before hospital discharge assessed mean diffusivity (MD) and fractional anisotropy (FA) in the cingulate, anterior thalamic radiations (ATR), and uncinate fasciculus. Hierarchical regression models examined associations between skin-to-skin care and white matter microstructure, adjusting for gestational age, health acuity, postmenstrual age at scan, and MRI coil type. Sensitivity analyses controlled for socioeconomic status and NICU visitation frequency.

**Results:** Skin-to-skin care duration was positively associated with MD in the cingulate (B = 0.002, p = 0.016) and ATR (B = 0.002, p = 0.020). Skin-to-skin care rate was also positively linked to MD in the ATR (B = 0.040, p = 0.041). Skin-to-skin care duration and rate were not associated with FA in the cingulate but skin-to-skin duration and rate were negatively associated with FA in the ATR (duration: B =-0.001, *p* = 0.020; rate: B =-0.017, p = 0.008). No significant associations were found for the uncinate fasciculus. Findings remained robust after adjusting for socioeconomic status and visitation frequency.

**Discussion:** This study provides novel evidence linking in-hospital experiences of skin-to-skin care to neonatal white matter development. These findings have important implications for understanding how family-centered neuroprotective practices, such as skin-to-skin care, may affect brain development to improve long-term developmental outcomes.

## Introduction

Preterm birth is a leading cause of neurodevelopmental disability worldwide.^1^ Approximately 50-67% of infants born very preterm (<32 weeks gestational age) experience neurodevelopmental impairments.^2^ Volumetric and diffusion magnetic resonance imaging studies link neurodevelopmental outcomes in children born very preterm to alterations neonatal white matter.^3–5^ White matter alterations are presumed to reflect disturbances in white matter maturation from hypoxia, ischemia, or inflammation induced by health complications associated with preterm birth.^6^ However, alterations in neonatal white matter may also reflect development differences relating to experiences and/or care practices in the neonatal intensive care unit (NICU) environment.^6,7^ Determining the impact of NICU caregiving experiences on neonatal white matter development has important implications for understanding how to modify neonatal care practices to support brain development and mitigate unfavorable long-term developmental outcomes.

Skin-to-skin care, also known as kangaroo care or kangaroo mother care (when provided by the mother), is a caregiving experience in which an infant, wearing only a diaper, is held to a caregiver’s bare chest.^8^ Skin-to-skin care is associated with multiple health benefits (e.g., improved cardiorespiratory stability, sleep, growth, and reduced infections, pain, and stress) and positive caregiving behaviors (e.g., increased bonding and breastfeeding), all likely to be important for brain development.^9–15^ Recent evidence has shown that in-hospital experiences of skin-to-skin care significantly account for cognitive outcomes beyond other well-known predictors of long-term outcomes, including family socio-economic background, infant health, and gestational age at birth.^16^ However, evidence explicitly linking skin-to-skin care to direct measures of neonatal structural brain development remains limited. Such data are likely to provide important insights into the neurobiological and/or neurophysiological processes that may be affected by skin-to-skin care experiences and that may, in turn, contribute to long-term outcomes.

In the present study, we examined the relations between measures of skin-to-skin care experienced by very preterm infants (<32 weeks gestational age) during their entire hospital stays and measures of white matter brain connectivity from diffusion MRI scans obtained just prior to hospital discharge. We hypothesized that variations in the amount of skin-to-skin care experienced by preterm infants would be associated with measures of white matter connectivity, assessed using two metrics from diffusion MRI: mean diffusivity (MD) and fractional anisotropy (FA). We based this hypothesis on data from both randomized controlled trials and observational studies in which higher amounts of skin-to-skin care were associated with significant improvements in infant neurobehavior, feeding, and stress. Additionally, white matter is sensitive to early experiences broadly.^10,17–21^ We focused our analyses on frontal and limbic pathways, including the cingulate, anterior thalamic radiations, and the uncinate fasciculus, because these white matter tracts functionally connect brain regions implicated in stress-regulation and socio-emotional development, processes that may be physiologically regulated by affective touch and tactile stimulation that occur during skin-to-skin care.^22–24^

## Method

### Design

All data, including MRI scans, were collected as part of standard of care at our hospital center. Instances and duration of parent-administered skin-to-skin care were charted in real time by clinical staff, primarily nurses, per unit protocols. These data were then collected from the Electronic Medical Record (EMR). This study was considered a minimal-risk retrospective chart review, therefore participants were not required to give consent. The experimental protocol was approved by Stanford University Institutional Review Board (#IRB-44480).

### Sample

Participants (*n=*86; 50% Female) were infants born less than 32 weeks gestational age between 5/1/2018 and 3/8/2020. Infants were included in analyses if (1) they were born in our center or admitted within the first 7 days of life so that we captured the majority of skin-to-skin experiences throughout their NICU stay and (2) they received MRI scanning at near-term age and prior to hospital discharge, per unit protocols. We excluded infants if (1) they were diagnosed with a genetic or congenital anomaly known to affect neurodevelopment (*n*=5), (2) had a diagnosis of grade 4 intraventricular hemorrhage which may interfere with analysis of MRI scans (IVH, *n*=8), or (3) were scanned at a postmenstrual age greater than 45 weeks (*n*=9).

Additional infants (*n*=15) were excluded from analyses due to poor quality MRI data determined by visual inspection, poor quality tracking in any of our tracts of interest (*n*=15), and/or scanned using a different head coil than all other participants (*n* = 1). See Figure 1 for consort diagram.

**Figure 1.**
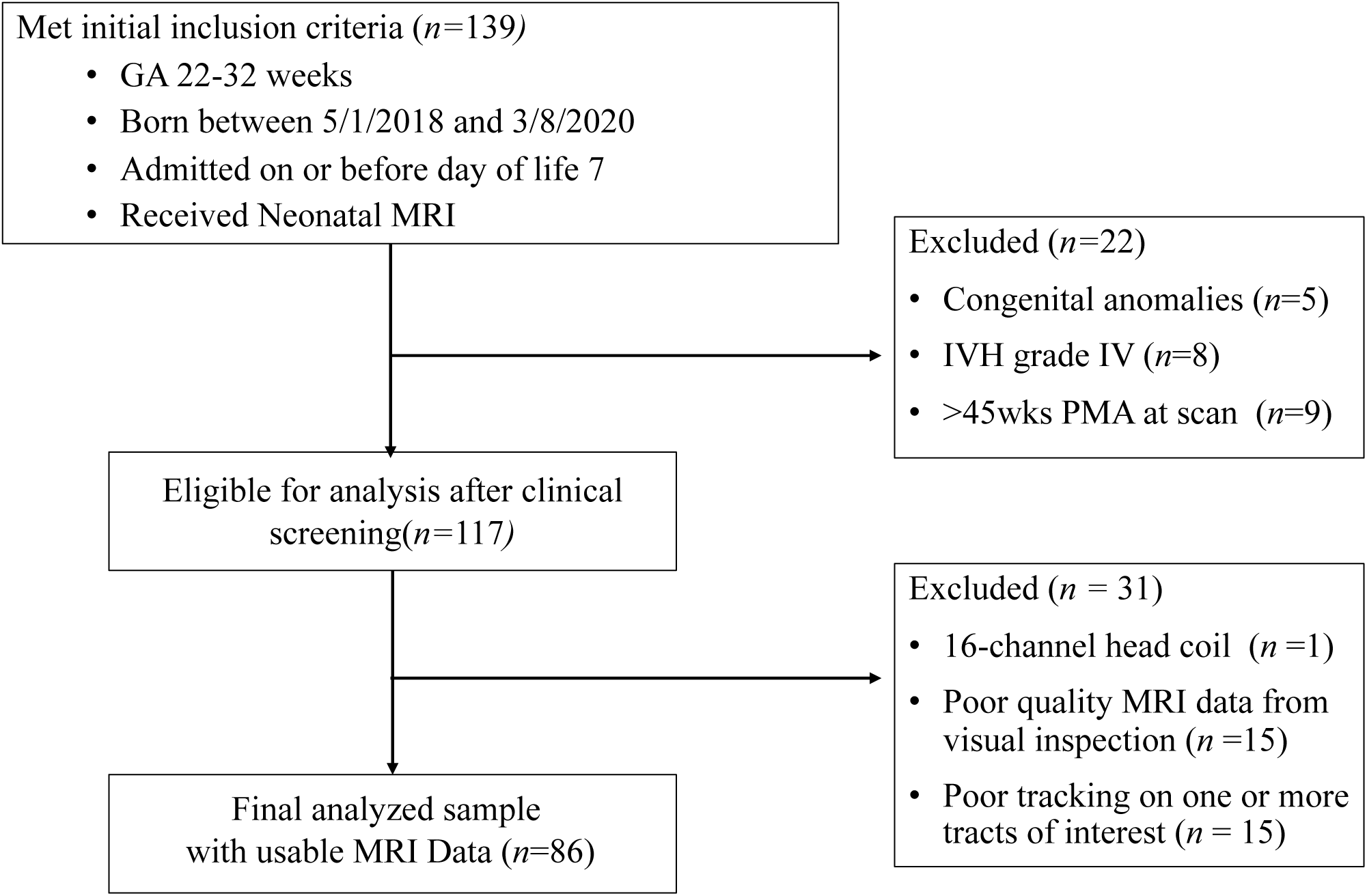
Consort diagram of eligible and included infants.

### Measures

#### Clinical and demographic measures

Clinical and demographic characteristics of all infants were extracted from the EMR, including gestational age at birth (weeks), weight at birth (kg), sex assigned at birth (male or female), length of hospital stay (days), and postmenstrual age at MRI (days). SES was indexed as public vs private insurance status. Since qualification for public insurance in California considers the income to needs ratio of the family, our metric for SES is thus reflective of income, as opposed to education, which was not reported in the EMR. We generated a binary health acuity score to categorize infants into those with none versus one or more of the following major comorbidities of prematurity: Bronchopulmonary dysplasia (BPD), Sepsis, Intraventricular Hemorrhage (IVH), and Necrotizing enterocolitis (NEC). BPD was defined as treatment with supplemental oxygen at 36 weeks postmenstrual age.^12^ Sepsis was defined as a positive blood culture or >7 days of antibiotics. IVH was defined as the presence of a grade I-III hemorrhage using the Papile classification system.^25^ NEC was defined as a diagnosis of medical or surgical NEC.

#### In-hospital skin-to-skin care and family visitation

Our NICU has developed a robust and comprehensive protocol for documenting developmental care activity and family visitation as they occur.^15,25–27^ Nurses chart developmental care type (skin-to-skin care, swaddled holding, touch, massage, music, talking, and singing), duration in minutes, and who was involved (mother, father, other family member, nurse, other staff member, or any combination of these people). Beginning in May 2018, our unit implemented a standardized developmental care pathway known as iRainbow, which provided clear clinical guidelines for determining infant eligibility for skin-to-skin care.^30^ This protocol ensured that decisions about skin-to-skin care were consistent across providers and based on defined medical criteria. We calculated skin-to-skin duration as the total number of minutes of family-administered skin-to-skin care prior to MRI divided by total number of instances of family-administered skin-to-skin care prior to MRI. We calculated skin-to-skin rate as the total number of minutes of family-administered skin-to-skin care prior to MRI divided by total days of hospital stay prior to MRI. We calculated skin-to-skin frequency as the total number of instances of family-administered skin-to-skin care prior to MRI divided by total days of hospital stay prior to MRI. We also calculated family visitation frequency as the total number of visitation instances prior to MRI divided by total days of hospital stay prior to MRI.

### Clinical MRI scanning and sequence parameters

During the study period, MRI scans were conducted prior to hospital discharge for all infants born very preterm (<32 weeks GA) in our NICU. Per clinical procedures, infants were scanned once stable in an open crib, requiring minimal respiratory support (no more than low-flow supplemental oxygen), and had reached a minimum postmenstrual age of 34 weeks. Before scanning, infants were fed, swaddled, and fitted with adhesive foam noise dampeners over their ears to block sounds from the scanner. Scans occurred during natural sleep with no sedation.

MRI scans were acquired using one of two 3.0T scanners (GE Discovery MR750 or GE Signa Premier) equipped with either an 8-channel HD head coil (n = 46) or a 32-channel HD head coil (n = 55) (General Electric Healthcare, Little Chalfont, UK). Sequences included in the clinical protocol and analyzed in this study included a high-resolution T1-weighted anatomical scan and 60-direction diffusion MRI scan. High-resolution T1-weighted scans were collected at ∼1mm^3^ spatial resolution and used as infant-specific anatomical reference for diffusion tractography analyses. Diffusion MRI included a high-angular resolution (60-direction) sequence with a b-value= 700sec/mm^2^ collected with a multi-slice echoplanar imaging (EPI) protocol for rapid image acquisition (∼3 min each, 6 min total) with two - six volumes at b=0. Diffusion MRI data were collected at 2.0mm^3^ spatial resolution (2.0mm x 2.0mm x 2.0mm isotropic voxels).

### Neuroimaging pre-processing and tractography analysis

MRI data were managed and analyzed using a cloud-based neuroinformatics platform (Flywheel.io). Procedures for dMRI pre-processing and tractography analyses of clinical neonatal scans are described in previous publications^31,32^ and are provided in detail in supplemental information. Using these procedures, we obtained measures of mean diffusivity (MD) and fractional anisotropy (FA) from each of the three pathways of interest. Figure 2a shows the resulting (left hemisphere) tractograms of the cingulate, anterior thalamic radiations, and the uncinate fasciculus from a single neonate displayed on their T1w image.

**Figure 2.**
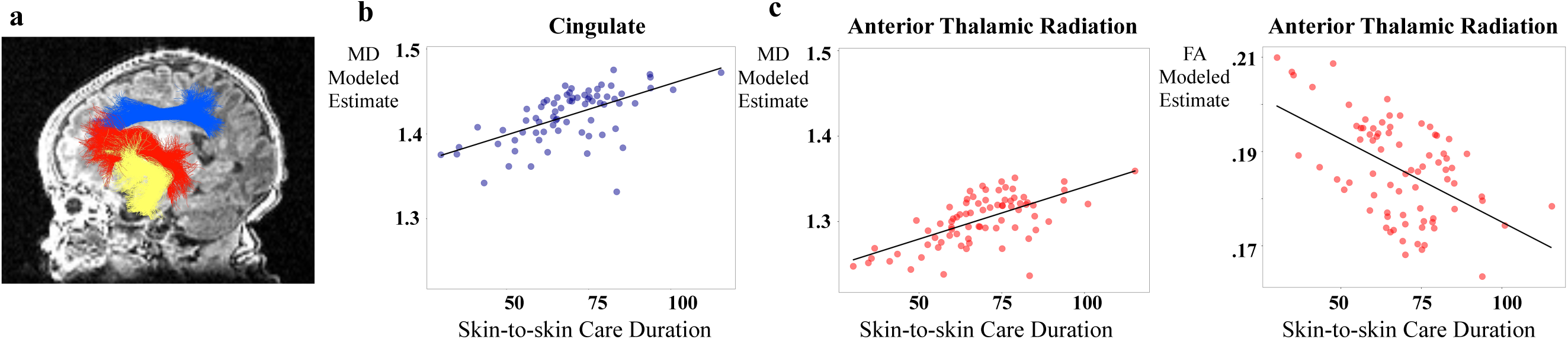
Associations between skin-to-skin care duration and white matter microstructure in preterm infants. Panel (a) illustrates a representative diffusion MRI image with tract reconstructions of the ATR (red), cingulum (blue), and uncinate (yellow). Panel (b) depicts the modeled association between skin-to-skin care duration and mean diffusivity (MD) in the cingulate (Table 2, Model 2). Panel c depicts the modeled association between skin-to-skin care duration and mean diffusivity (MD, Table 3, Model 2) and fractional anisotropy (FA, Table 3, Model 5) in the ATR.

#### Statistical Analysis

All analyses were conducted using R version 4.1.2. We examined descriptive statistics for visitation frequency, each metric of skin-to-skin care, and white matter microstructure measures. All variables were inspected for outliers and tested for normality. Any variable with a value above or below 3 standard deviations (SDs) from the mean was winsorized. Shapiro-Wilk tests identified skin-to-skin care rate and frequency as non-normal, so they were transformed using a base 10 log. To reduce the number of comparisons, we collapsed left and right tracts into bilateral metrics. All left-right tract pairs were significantly correlated (*r*s between 0.41 and 0.57 for MD; *r*s between 0.36 and 0.63 for FA; all *p*s < 0.01).

Next, we assessed correlations among our skin-to-skin care metrics. Skin-to-skin care duration, rate, and frequency were all significantly correlated. Notably, rate and frequency were very highly correlated (*r* = 0.92, *p* < 0.001), while duration showed more moderate correlations with both rate (*r* = 0.57, *p* < 0.001) and frequency (*r* = 0.37, *p* < 0.001). To reduce redundancy in our analyses, we chose to focus on duration and rate as our primary predictors of interest.

Zero-order correlations inspected relations between predictors and white matter microstructure measures. For each white matter tract (cingulate, anterior thalamic radiations, and uncinate fasciculus), we ran two sets of hierarchical linear regression models, one with MD as the outcome variable, and another with FA as the outcome variable. In each set, we first included only the covariates (gestational age at birth, health acuity score, postmenstrual age at scan, coil type), and then added skin-to-skin care duration or rate as predictors. This approach allowed us to assess the unique contribution of each skin-to-skin care metric to white matter microstructure while controlling for key clinical and scan-related factors that might impact brain measurement outcomes.^32,33^

We performed sensitivity analyses to test the robustness of these findings and to account for additional potential confounders. Specifically, we ran additional models that included socioeconomic status (SES) and visitation frequency as covariates. We included SES because it is known to be related to skin-to-skin care engagement,^27,29^ brain development,^34^ and neurodevelopmental outcomes.^29,34,35^ Including visitation frequency allowed us to understand the specificity of skin-to-skin care effects beyond variation in general family involvement. These analyses help to determine whether the observed relationships between skin-to-skin care and white matter microstructure persisted when considering important social factors. All significance levels were set at *p <* 0.05.

## Results

### Sample Characteristics

As shown in Table 1, the sample consisted of 86 very preterm infants (50% female), born at 29 weeks gestational age and weighing 1200 grams, on average. Approximately 47% of the infants had public insurance, serving as a proxy for lower socioeconomic status. Infants were hospitalized for an average of 2 months, though one infant had a length of stay of more than 5 months. 40% of the infants experienced one or more of the major comorbidities of prematurity.

**Table 1.** Descriptive Statistics of the Sample (*n*=86)

|  | **M (SD) or n (%)** | **Min** | **Max** |
| --- | --- | --- | --- |
| Female (%) | 43 (50.0%) | - | - |
| GA at Birth (weeks) | 28.96 (2.23) | 22.7 | 31.8 |
| Birthweight (g) | 1176.29 (343.27) | 530.00 | 1870.00 |
| Public Insurance (%)<sup>1</sup> | 40 (46.5%) | - | - |
| Length of Stay (days) | 63.34 (29.42) | 19.78 | 177.14 |
| Health Acuity (%)<sup>2</sup> | 31 (36.0%) | - | - |
| BPD (%) | 15 (17.4%) | - | - |
| IVH (%) | 13 (15.1%) | - | - |
| NEC (%) | 10 (11.6%) | - | - |
| Sepsis (%) | 6 (7.0%) | - | - |
| Visitation Frequency<sup>3</sup> | 0.93 (0.38) | 0.23 | 2.01 |
| Skin-to-skin Care Duration<sup>4</sup> | 68.72 (15.56) | 30.00 | 115.41 |
| Skin-to-skin Care Rate<sup>5</sup> | 23.67 (20.11) | 0.00 | 85.64 |
| Skin-to-skin Care Frequency<sup>6</sup> | 0.32 (0.25) | 0.00 | 1.08 |
| PMA at MRI (weeks) | 36.53 (1.57) | 34.43 | 43.86 |
| 32 Channel Head Coil | 48 (55.8%) | - | - |
| MD Cingulate | 1.42 (0.08) | 1.21 | 1.65 |
| FA Cingulate | 0.16 (0.02) | 0.10 | 0.23 |
| MD Anterior Thalamic Radiation | 1.30 (0.08) | 1.14 | 1.58 |
| FA Anterior Thalamic Radiation | 0.19 (0.03) | 0.12 | 0.28 |
| MD Uncinate | 1.38 (0.09) | 1.11 | 1.63 |
| FA Uncinate | 0.17 (0.03) | 0.09 | 0.24 |

<sup>1</sup> SES as defined by the percentage of families with public vs. private health insurance.
<sup>2</sup> Health acuity score reflecting diagnoses of one or more of the following conditions: bronchopulmonary dysplasia (BPD), intraventricular hemorrhage (IVH), necrotizing enterocolitis (NEC), sepsis.
<sup>3</sup> Number of instances of family visitation/number of days of inpatient hospital stay.
<sup>4</sup> Number of minutes of skin-to-skin care/ number of instances of skin-to-skin care.
<sup>5</sup> Number of minutes of skin-to-skin care/number of days of inpatient hospital stay.
<sup>6</sup> Number of instances of skin-to-skin care/number of days of inpatient hospital stay.

Families in our study varied in amounts of hospital visitation and skin-to-skin care engagement. On average, families visited the NICU approximately once per day, with some visiting more than twice daily. When families engaged in skin-to-skin care, they tended to do so for around 70 minutes, on average, ranging from about 30 minutes to nearly 2 hours per instance. Throughout their infant’s hospitalization, families engaged in skin-to-skin care at an average daily rate of about 24 minutes/day. However, there was substantial variability in this practice; 10 infants (11.6%) had families providing more than 50 minutes of daily skin-to-skin care, while 6 infants (7.0%) had families who did not engage in any skin-to-skin care during their hospitalization. Overall, families engaged in skin-to-skin care at frequencies of around 2-3 days/week. Infants underwent MRI scans at around 36.5 weeks, with about 50% being scanned with 32 channel head coils. Mean MD and FA for each of the three pathways examined are reported in Table 1.

Zero-order correlations revealed that skin-to-skin care duration was significantly associated with MD of the cingulate (*r* = 0.24, *p* = 0.046) and marginally associated with MD (*r* = 0.22, *p* = 0.050) and FA (*r* =-0.20, *p* = 0.078) of the ATR. Skin-to-skin care rate was significantly associated with MD of the cingulate (*r* = 0.23, *p* = 0.046).

### Hypothesis Testing Cingulate

Table 2 documents the unique contribution of skin-to-skin care duration and rate to MD and FA of the Cingulate. Models 1-3 show associations between predictors and Cingulate MD. In Model 1, covariates (gestational age, health acuity, postmenstrual age at scan, and coil type) accounted for approximately 8% of the variance in Cingulate MD. Model 2 demonstrates that skin-to-skin care duration was significantly associated with MD (B = 0.002, *p* = 0.016), conferring 7.6% additional variance over and above covariates. Model 3 demonstrates that skin-to-skin rate was not significantly associated with Cingulate MD over and above covariates.

**Table 2.**
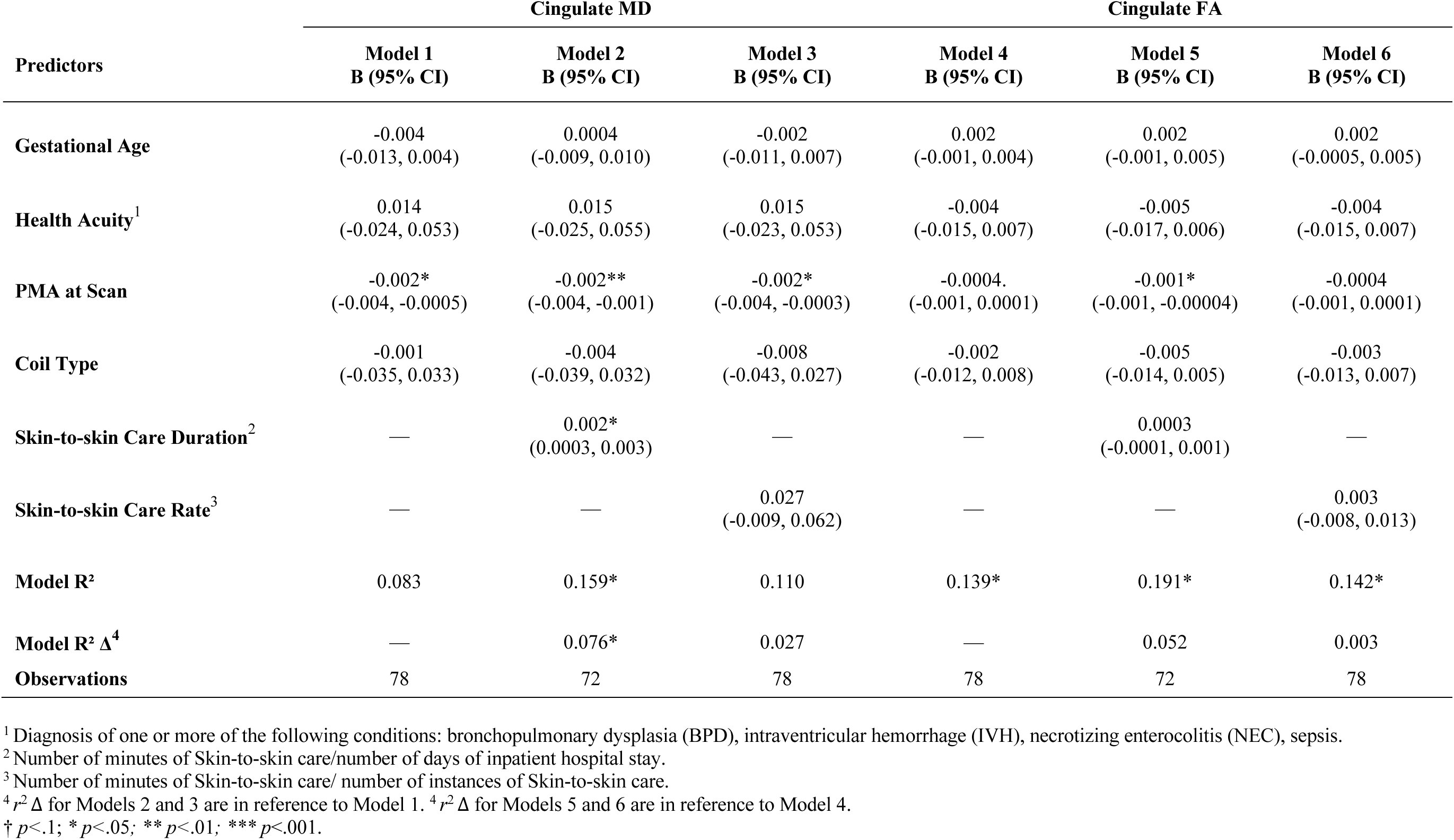
Multiple regression models (unstandardized coefficients) predicting MD and FA of the bilateral Cingulate.

Models 4-6 show associations between predictors and Cingulate FA. In Model 4 covariates (gestational age, health acuity, postmenstrual age at scan, and coil type) accounted for approximately 14% of the variance in Cingulate FA. Models 5 and 6 demonstrate that neither skin-to-skin duration nor rate predicted Cingulate FA over and above covariates. Figure 2b depicts the modeled association between skin-to-skin care duration and MD of the cingulate controlling for covariates.

### Anterior Thalamic Radiations (ATR)

Table 3 documents the unique contribution of skin-to-skin care duration and rate to MD and FA of the Anterior Thalamic Radiations (ATR). Models 1-3 show associations between predictors and ATR MD. In Model 1, covariates (gestational age, health acuity, postmenstrual age at scan, and coil type) accounted for approximately 6% of the variance in ATR MD. Model 2 demonstrates that skin-to-skin care duration was significantly associated with MD (B = 0.002, *p* = 0.02 conferring 5.8% additional variance over and above covariates. Model 3 shows that skin-to-skin care rate was significantly associated with ATR MD (B = 0.040, *p* = 0.041), explaining 5.0% additional variance. Models 4-6 show associations between predictors and ATR FA. In Model 4 covariates (gestational age, health acuity, postmenstrual age at scan, and coil type) accounted for approximately 10% of the variance in ATR FA. Model 5 demonstrates that skin-to-skin care duration was significantly negatively associated with ATR FA (B =-0.001, *p* = 0.020), accounting for 5.2% additional variance beyond covariates. Model 6 demonstrates that skin-to-skin care rate was significantly negatively associated with ATR FA (B =-0.017, *p* = 0.008) contributing 7.8% additional variance over covariates. Figure 2c depicts the modeled association between skin-to-skin care duration and MD and FA of the ATR controlling for covariates.

**Table 3.**
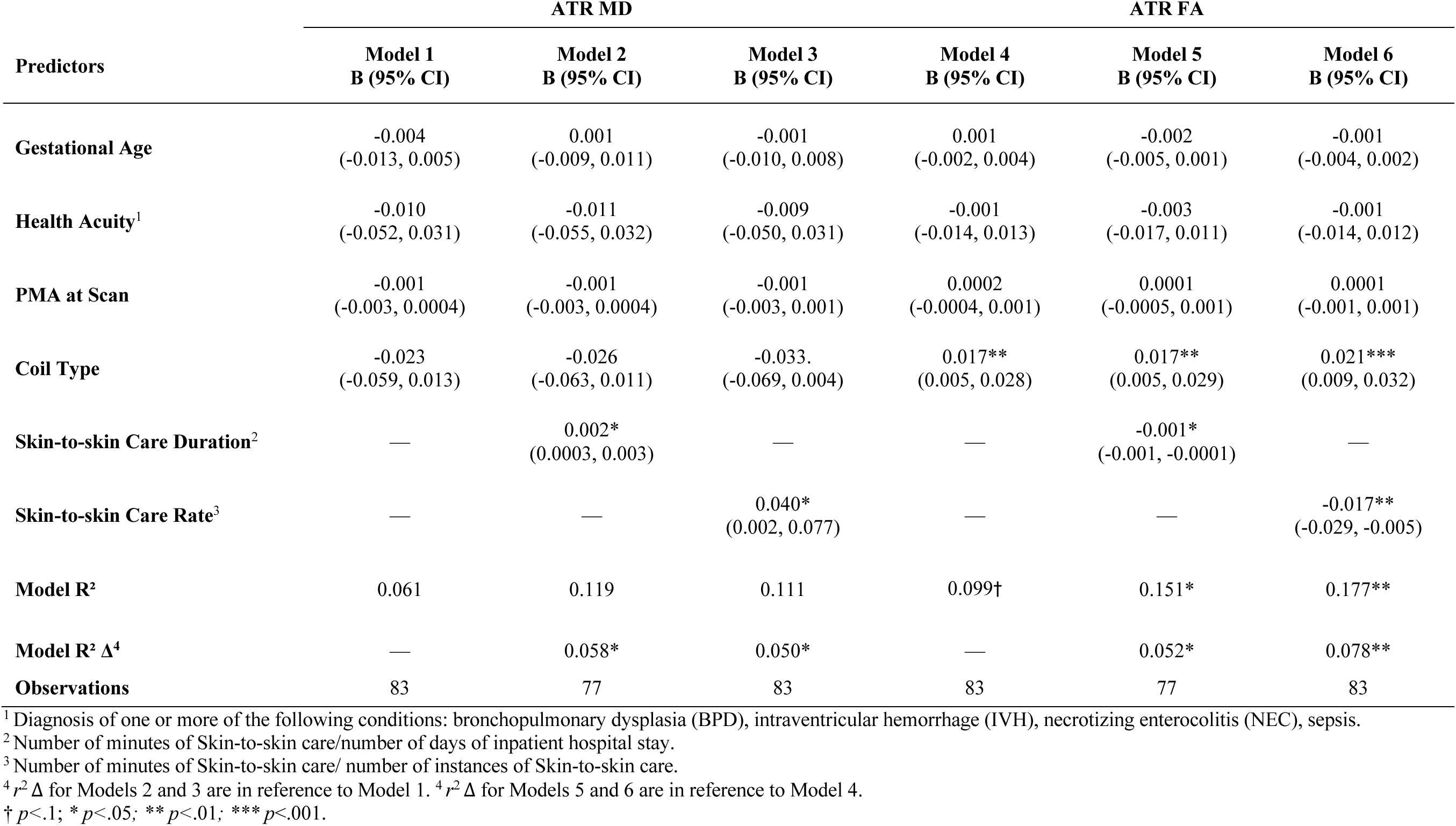
Multiple regression models (unstandardized coefficients) predicting MD and FA of the bilateral Anterior Thalamic Radiations (ATR).

| Predictors | ATR MD |  |  | ATR FA |  |  |
| --- | --- | --- | --- | --- | --- | --- |
|  | Model 1<br>B (95% CI) | Model 2<br>B (95% CI) | Model 3<br>B (95% CI) | Model 4<br>B (95% CI) | Model 5<br>B (95% CI) | Model 6<br>B (95% CI) |
| **Gestational Age** | -0.004<br>(-0.013, 0.005) | 0.001<br>(-0.009, 0.011) | -0.001<br>(-0.010, 0.008) | 0.001<br>(-0.002, 0.004) | -0.002<br>(-0.005, 0.001) | -0.001<br>(-0.004, 0.002) |
| **Health Acuity<sup>1</sup>** | -0.010<br>(-0.052, 0.031) | -0.011<br>(-0.055, 0.032) | -0.009<br>(-0.050, 0.031) | -0.001<br>(-0.014, 0.013) | -0.003<br>(-0.017, 0.011) | -0.001<br>(-0.014, 0.012) |
| **PMA at Scan** | -0.001<br>(-0.003, 0.0004) | -0.001<br>(-0.003, 0.0004) | -0.001<br>(-0.003, 0.001) | 0.0002<br>(-0.0004, 0.001) | 0.0001<br>(-0.0005, 0.001) | 0.0001<br>(-0.001, 0.001) |
| **Coil Type** | -0.023<br>(-0.059, 0.013) | -0.026<br>(-0.063, 0.011) | -0.033<br>(-0.069, 0.004) | 0.017**<br>(0.005, 0.028) | 0.017**<br>(0.005, 0.029) | 0.021***<br>(0.009, 0.032) |
| **Skin-to-skin Care Duration<sup>2</sup>** | — | 0.002*<br>(0.0003, 0.003) | — | — | -0.001*<br>(-0.001, -0.0001) | — |
| **Skin-to-skin Care Rate<sup>3</sup>** | — | — | 0.040*<br>(0.002, 0.077) | — | — | -0.017**<br>(-0.029, -0.005) |
| **Model R<sup>2</sup>** | 0.061 | 0.119 | 0.111 | 0.099† | 0.151* | 0.177** |
| **Model R<sup>2</sup> Δ<sup>4</sup>** | — | 0.058* | 0.050* | — | 0.052* | 0.078** |
| **Observations** | 83 | 77 | 83 | 83 | 77 | 83 |

<sup>1</sup> Diagnosis of one or more of the following conditions: bronchopulmonary dysplasia (BPD), intraventricular hemorrhage (IVH), necrotizing enterocolitis (NEC), sepsis.
<sup>2</sup> Number of minutes of Skin-to-skin care/number of days of inpatient hospital stay.
<sup>3</sup> Number of minutes of Skin-to-skin care/ number of instances of Skin-to-skin care.
<sup>4</sup> $r^2 \Delta$ for Models 2 and 3 are in reference to Model 1. <sup>4</sup> $r^2 \Delta$ for Models 5 and 6 are in reference to Model 4.
† $p < .1$ ; \* $p < .05$ ; \*\* $p < .01$ ; \*\*\* $p < .001$ .

**Table 4.**
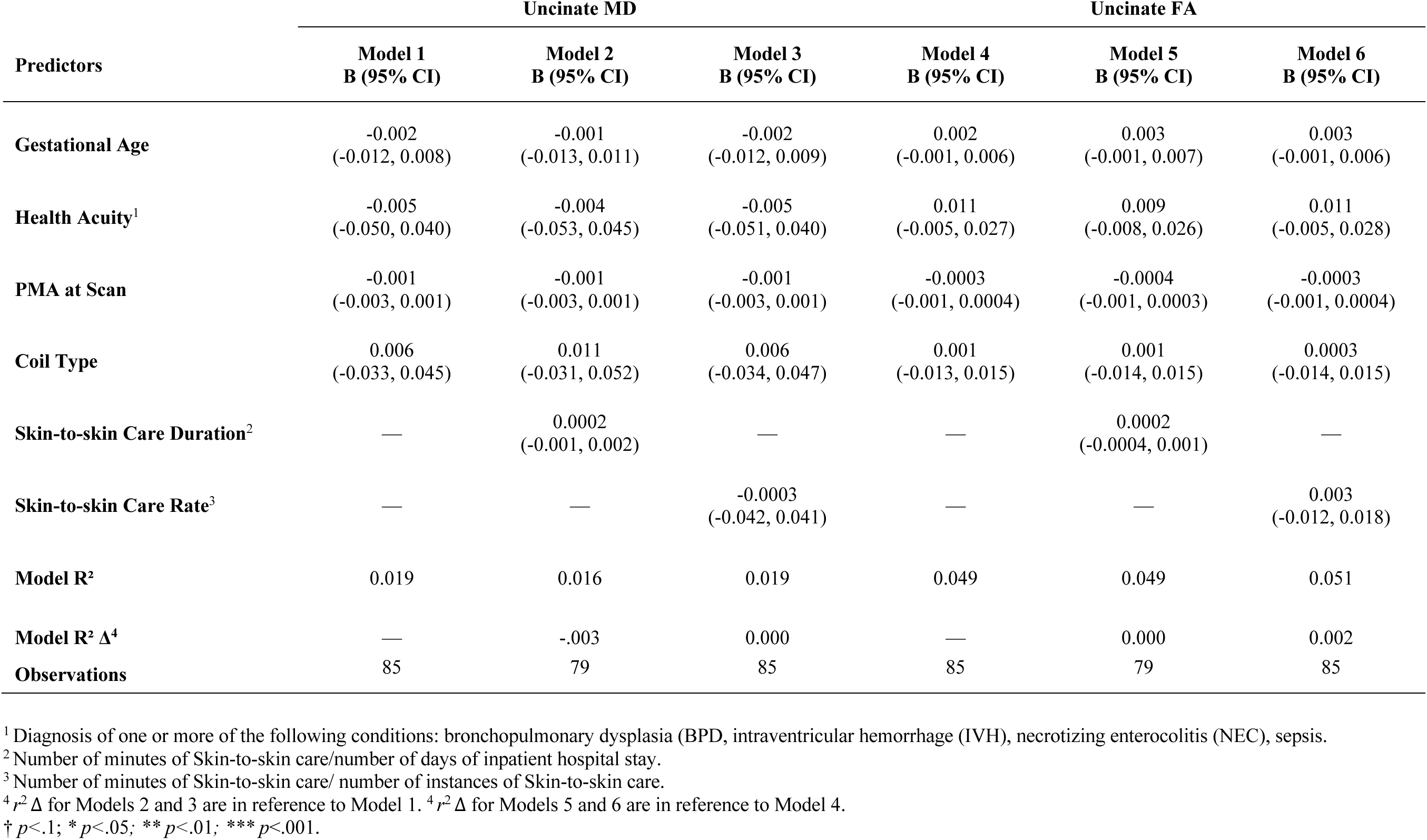
Multiple regression models (unstandardized coefficients) predicting MD and FA of the bilateral Uncinate.

### Uncinate Fasciculus

Zero-order correlations did not reveal any significant relationships between skin-to-skin care metrics and MD or FA of the uncinate fasciculus. Neither skin-to-skin care duration nor rate was significantly associated with MD or FA in the uncinate fasciculus in regression analyses (all *p*s > 0.1).

### Sensitivity Analyses

We conducted sensitivity analyses by including visitation frequency and socioeconomic status (SES) as additional covariates in our models. The pattern of results remained largely unchanged. For the cingulate, skin-to-skin care duration remained significantly associated with MD (B = 0.002, *p* = 0.018). For the ATR, skin-to-skin care duration remained significantly associated with MD (B = 0.002, *p* = 0.023) and FA (B =-0.0005, p = 0.036) but associations with rate became marginal (MD: B = 0.050, p = 0.056; FA: B =-0.015, p = 0.078). As in the primary analyses, no significant associations were found for the uncinate fasciculus.

## Discussion

Here, we provide evidence linking in-hospital experiences of skin-to-skin care to measures of neonatal structural brain development in infants born very preterm. Specifically, we found that the duration and rate of skin-to-skin care experiences during NICU hospitalization were significantly associated with measures of white matter microstructure obtained from two frontal and limbic white matter pathways: the cingulate and anterior thalamic radiations.

Importantly, these associations remained significant after accounting for clinical, developmental, socio-demographic, and scan-related factors shown previously to be related to neonatal white matter development.^32,34–36^ Taken together, these findings provide evidence that early family-centered caregiving experiences in the NICU may be an important and unique contributor to neonatal brain development in preterm born infants.

Prior evidence linking skin-to-skin care experiences to brain development has either come from studies assessing the safety of skin-to-skin care using indirect physiological markers of neural functioning (e.g., heart rate variability, oxygen saturation) or randomized trials that have assessed structural and functional brain development in older children and adults who had experienced skin-to-skin care as infants.^18,37–40^ Our study provides novel evidence that skin-to-skin care may influence brain development as early as the neonatal period, even before infants are discharged from the hospital. This extends prior work by demonstrating that the effects of skin-to-skin care on the brain are not only long-term and potentially mediated by enhanced parent-infant bonding, but also more immediate and proximal to the experience itself. Our findings are complementary to prior work that has linked white matter development to adverse painful experiences in the neonatal period, such as medical procedures requiring skin breaks.^41^ Taken together, these findings add to growing evidence that white matter development is sensitive to experience and is likely to be impacted by NICU caregiving activities, including those presumed to be neuroprotective or aversive. Further research is needed to determine if optimizing neuroprotective activities can mitigate the negative effects of medical complications and necessary clinical procedures that may induce pain on neonatal brain development and longer-term outcomes.

There are multiple plausible mechanisms through which skin-to-skin might influence neonatal brain development. Skin-to-skin care has been associated with reduced physiological stress, improved weight gain, enhanced cardiorespiratory stability, and better sleep, all of which could directly or indirectly contribute to white matter properties and its development.^10,12,19,39,40,42^ Skin-to-skin care might also impact white matter development by providing direct multi-sensory stimulation to tactile, olfactory, auditory, and vestibular systems. These mechanisms offer a compelling explanation for how skin-to-skin care might support brain development, yet the correlational nature of this study means we cannot exclude the possibility that within-infant characteristics – including white matter microstructure - may have led to a higher tolerance for prolonged sessions of skin-to-skin care. However, because our analyses controlled for gestational age and infant health status, we do not believe that our findings simply reflect healthier infants receiving more care. Future studies using randomized interventions or longitudinal designs are needed to establish causality and further elucidate the extent to which skin-to-skin actively contributes to neonatal brain maturation.

We observed slightly stronger associations between white matter development and skin-to-skin care duration compared to rate. This finding may suggest that the length of individual skin-to-skin sessions, rather than the total accumulated time across hospitalization, is particularly important for supporting brain characteristics. Other studies have shown particular benefits of skin-to-skin duration on cardio-respiratory and nutritional outcomes.^43,44^ Longer, sustained periods of skin-to-skin care may provide more consistent physiological regulation and better support for sleep, enabling neurobiological processes that influence white matter organization.

Additionally, it is notable that many infants in our study received average skin-to-skin durations at or below the World Health Organization’s recommended guideline of 60 minutes per session. However, we still detected associations with white matter development.^8^ This finding suggests that even relatively low levels of skin-to-skin care may influence neonatal brain structure, reinforcing the need to explore optimal skin-to-skin care dosing in future studies.

The direction of the association between skin-to-skin care and mean white matter metrics and limited associations with fractional anisotropy (FA) provide important insights for how early life experiences may impact neonatal brain development. During development, MD negatively correlates with age^45^ and may reflect reductions in water content from increases in tissue density, including those from myelination.^46^ Here, the positive association between MD and skin-to-skin care suggests that tissue properties, separate from those that contribute to reductions in MD over development, are correlated with skin-to-skin contact. We speculate that skin-to-skin contact could limit neuroinflammatory responses related to comorbidities of preterm birth (e.g., bronchopulmonary dysplasia, hypothalamic-pituitary-axis immaturity) or extrinsic stressors (e.g., pain, lack of parental contact/nutritive care). Reductions in inflammatory responses induced by reactive species (e.g., microglia) could alter tissue properties that may increase MD^47^ and/or slow cellular aging, a proposed mechanism through which environmental stress may affect brain development.^48^ FA is highly sensitive to fiber coherence and is commonly associated with within-participant behavioral skills, whereas MD has been associated with behavioral interventions.^49,50^ Fiber coherence may thus not be as sensitive to caregiving experiences. More research, including additional MRI protocols or histological validation, is needed to understand the balance of tissue properties that contribute to diffusion metrics in the immature neonatal brain and their sensitivity to environmental factors.

This study has limitations, but several methodological strengths help mitigate concerns.

While our measures of skin-to-skin care were extracted from the electronic medical record, which may introduce variability in documentation, the charting of developmental care in our NICU is highly reliable. Clinical staff are routinely trained to document skin-to-skin care with the same attention to detail as they use for documenting nutrition, elimination, weight, and other clinical data. In many ways, charting by hospital personnel may be more accurate than the method used in most prior studies, e.g., parent-reported skin-to-skin care journals, because our data are derived from an objective, observational source and less susceptible to social desirability bias. Furthermore, the presence of a standardized clinical care pathway for determining eligibility for skin-to-skin care reduces provider variability in offering this intervention and may promote more equitable delivery across infants with similar medical profiles.

Beyond these considerations, several limitations should be noted. First, while we quantified skin-to-skin care based on duration and rate, we did not capture qualitative aspects of these interactions, such as caregiver engagement, positioning, or the emotional context of the experience. These factors may be important for understanding the full impact of skin-to-skin care on brain development. Second, our study was conducted within a single high-resourced NICU with a well-established system for charting developmental care, which may limit generalizability to other NICU settings with different resources, cultural practices, or documentation protocols.

Third, our sample included only infants who remained stable enough to undergo MRI prior to discharge, potentially excluding the most critically ill infants who may have different patterns of skin-to-skin care engagement and neurodevelopmental trajectories.

## Conclusion

The mean duration per instance of skin-to-skin contact and the mean number of minutes of skin-to-skin per hospital day were associated with measures of white matter microstructure in children born very preterm. These findings suggest that skin-to-skin contact may be neuroprotective. Future research should identify mechanisms of this effect, which may result from reductions in inflammatory responses or slow cellular aging, based on the direction of association. Future studies should further assess whether these early differences in brain microstructure translate to positive long-term cognitive, language, or socio-emotional outcomes and whether white matter microstructure may serve as a mediator between skin-to-skin care and neurodevelopmental outcomes.

## Data Availability

All data produced in the present study are available upon reasonable request to the authors

