## Supplemental Materials for "Skin-to-skin holding in relation to white matter connectivity in infants born preterm"

### Image Preprocessing

Procedures for dMRI analyses were implemented in Reproducible Tract Profiles (RTP) (https://github.com/vistalab/RTP-pipeline) .[17,18] Diffusion preprocessing, modeling and tractography within RTP is largely based on software from FSL (https://fsl.fmrib.ox.ac.uk/fsl/fslwiki), MRTrix3 (https://www.mrtrix.org/) and Automated Fiber Quantification[25] (AFQ, https://github.com/yeatmanlab/AFQ). A full list of software dependencies can be found here (https://github.com/vistalab/RTP-pipeline/wiki).

RTP consists of three main steps: (i) Structural processing and Region of Interest (ROI) creation (ii) dMRI preprocessing (RTP-preproc) and (iii) whole-brain tractography and tract segmentation (RTP-Pipeline). Detailed information for processing steps were described in detail by Lerma-Usabiaga et al.[17] and Liu et al.[18] We modified RTP to perform segmentation of the T1w image using Infant Freesurfer[19] (https://surfer.nmr.mgh.harvard.edu/fswiki/infantFS) to accommodate for the immature neonate brain. We substituted a neonatal template (Edinburgh Neonatal Atlas, ENA33)[20] with ROIs for tract identification and segmentation.

We performed the following diffusion image preprocessing steps (RTP-preproc): (i) data denoising with principal component analysis[21,22] and Gibbs ringing correction,[23] (ii) eddy current and motion correction,[24] (iii) and anatomical alignment of diffusion data to the average of the non-diffusion-weighted volumes, which were registered to the infant's high-resolution ac-pc aligned anatomical image using rigid body transformation.

### Diffusion Tractography

The preprocessed dMRI data output from RTP-preproc served as the input for diffusion metrics modeling, whole-brain tractography, and tract segmentation in RTP-pipeline. White matter diffusion tensor metrics were calculated based on the diffusion tensor model. Constrained spherical deconvolution model (CSD),[26] with eight spherical harmonics (lmax = 8) was used to calculate fiber orientation distributions (FOD) for each voxel. CSD tractography and FA masks were adjusted for the immaturity of the neonatal brain to have FA mask thresholds of 0.15 with FOD of 0.08. The CSD FODs were used for dMRI tractography, which consisted of (i) Ensemble Tractography[27] to estimate the whole-brain white matter connectome. MRtrix3 was used to generate three candidate connectomes that varied in their minimum angle parameters (50°, 30°, 10°).[28] For each candidate connectome, a probabilistic tracking algorithm (iFOD2) was used with a step size of 1 mm, a minimum length of 10 mm, a maximum length of 200 mm, and an FOD stopping criterion of 0.04. (ii) Spherical-deconvolution Informed Filtering of Tractograms to improve the quantitative nature of the ensemble connectome by filtering the data such that the streamline densities match the FOD lobe integral. The resulting ensemble connectome retained 500,000 streamlines. (iii) Concatenation of the three candidate connectomes into one ensemble connectome. (iv) AFQ[25] was used to segment and refine the resulting whole-brain connectome of each child into the tracts of interest: the left and right cingulate, anterior thalamic radiations and uncinate.[29,30] Figure XX shows the ROIS and resulting tracts on a T1w image from a representative participant born XX GA an imaged at XX weeks PMA. Mean tract mean diffusivity (MD) was calculated for the core of the tract between defining ROIs.
